# Altered functional connectivity relates to motor performance deficits in bipolar but not unipolar depression

**DOI:** 10.1101/2021.11.26.21266905

**Authors:** Lara E. Marten, Aditya Singh, Anna M. Muellen, Sören M. Noack, Vladislav Kozyrev, Renate Schweizer, Roberto Goya-Maldonado

## Abstract

Underpinnings of psychomotor deficits in bipolar and unipolar depression remain underexplored. Here, we hypothesize that motor performance deficits in patients may be partially explained by altered functional connectivities between hand primary motor cortex and posterior cingulate cortex with supplementary motor area.

95 participants between 18-65 years of age, including bipolar depressed, unipolar depressed, and sex-, age-, and education-matched healthy controls, participated in this observational study with two separate visits about five weeks apart, during which the patients received psychopharmacological treatment. Motor performance was measured with a finger-tapping-task and related to functional connectivity from individual seeds in hand primary motor cortex and posterior cingulate cortex as well as to the default mode and sensory motor networks from resting state functional MRI data.

79 participants (45.6% females, 21 bipolar depressed, 27 unipolar depressed and 31 healthy controls) were included in the analysis. Using a finger-tapping-task, the groups differed in motor performance (ANOVA factor “group” *F(2,76) = 4.122*; *p = 0.020*) and bipolar depressed but not unipolar depressed showed performance deficits compared to controls (post-hoc-test *p = 0.023* and *p = 0.158* respectively). Behavioral performance correlated with functional coupling of posterior cingulate cortex - supplementary motor area, but not with coupling of primary motor cortex - supplementary motor area at cluster-wise correction level *p FWEc < 0.05*. Correlation differences were seen in posterior cingulate cortex - supplementary motor area (healthy controls>bipolar depressed, unipolar depressed>bipolar depressed) at second visit and in primary motor cortex - supplementary motor area (bipolar depressed>unipolar depressed) at both visits at cluster-wise correction level *p FWEc<0.05*. Motor performance did not relate to functional coupling of sensory motor network - anterior (visit 1 *p = 0.375*, visit 2 *p = 0.700*) or - posterior (visit 1 *p = 0.903*, visit 2 *p = 0.772*) default mode network.

Motor performance deficits were seen exclusively in bipolar depressed and related to reduced posterior cingulate cortex - supplementary motor area functional connectivity at rest. Our results shed new light on a possible disruption in the anticorrelation between these regions, which seems fundamental for the preservation of motor skills. Given that nuisance factors were controlled for in the study, it is unlikely that the main results are biased by lefthanders, medication load, seed volumes, or differences in movements during MRI scanning. If these findings are confirmed, new targeted non-invasive interventions, such as repetitive transcranial magnetic stimulation, may be more effective against psychomotor deficits in bipolar depression, when aimed at modulating the supplementary motor area.

## 1. Introduction

Psychomotor functioning involves processes that range from planning, initiating and executing movements.^1,for review see 2^ Slower or inefficient information processing may derive from alterations in particular neural regions, or entire networks, that are essential to the circuitry.^3–7^ The precentral gyrus contains the primary motor cortex (M1), which is in charge of starting the execution loop of movements of the contralateral limb.^8,9^ For the coordinated adjustment of muscles implicated in the intended hand movement, the supplementary motor area (SMA) integrates signals of frontal planning with sensorial, proprioceptive and cognitive information from other brain regions and forwards it to the M1.^8–10,for a review see 11^ Another pivotal region involved in successful motor responses is the posterior cingulate cortex (PCC), a region belonging to the default mode network (DMN). PCC plays a role in integrating signals from somatosensory areas and dorsal visual stream via parietal cortical areas for spatial processing and action in space.^for review see 12^ In stroke patients with impairments in motor performance, strengthened resting state functional connectivity between the M1 and the PCC was found to be critical for improvements in motor performance.^17^ Studies have shown that the PCC is also involved in emotional processing and rumination of negative thoughts.^13–16^ This makes it an interesting target for research on depression, since psychomotor symptoms in neuropsychiatric disorders may involve behaviors arising from more than just the motor circuitry.^for review see 6^ To broaden the understanding of the contribution of M1 and PCC also in motor performance deficits in depression,^18–21^ studies that unveil their longitudinal functional connectivity are crucial.

Deficits in psychomotor functioning is a typical symptom cluster of major depressive disorder and the depressive phase of bipolar disorder^22,23^ and can make a decisive contribution to unsuccessful treatment^24,25^ as well as to social functioning.^26^ In spite of their impact on patient outcomes, their manifestation is often assumed to be either part to residual symptoms or medication side effects, leaving their neural correlates and development of potentially targeted treatments poorly explored. The identification of more objective differences involving motor performance and its neural correlates in groups of bipolar depressed (BD), unipolar depressed (UD) patients and healthy controls (HC) could support the development of novel biomarkers. Furthermore, such neurofunctional biomarkers of BD and UD can offer the chance to better understand the basis of motor symptom manifestation and treatment improvement,^for review see 27^ for example, by applying more targeted transcranial magnetic stimulation (TMS) protocols.^for review see 28^

Different paradigms can be used to assess and compare the motor performance of UD, BD, HC and recovered patients.^29–34^ In the present study, we have chosen the finger-tapping-task (FTT) paradigm, considering that it dismisses working memory,^35,36^ which allows clearer conclusions about the movement process itself. It has been previously shown that motor performance in FTT is impaired in BD,^18,19^ but not always in UD,^5,20,21,37–39^. This reinforces an importance of understanding the underlying brain mechanisms leading to such differences. In light of possible residual motor symptoms,^40,41^ the fundamental evaluation of motor performance, e.g. using the FTT, related to baseline M1 and PCC functional connectivity in depressed subjects undergoing treatment remains unexplored to our knowledge. Therefore, our aim with this study was to test whether resting state functional connectivity of M1 and PCC with SMA, which is accessible on the MRI scanner to all patients, regardless of how severely depressed they are, can explain behavioral differences in subjects. Functional connectivity between the right M1 and the SMA, an integrative center of the sensory motor network (SMN), can be a positive predictor of performance during the FTT in HC.^42^ Thus, baseline functional dysconnectivities with the SMA could potentially explain behavioral differences between BD and UD patients in contrast to HC.

Large-scale networks can differ according to mood states in bipolar disorder.^43–45^ A model involving the interaction between the SMN and the DMN has been recently suggested to explain psychomotor deficits in affective and many other disorders.^for review see 6^ In BD, it was proposed that the interaction between networks is imbalanced towards the DMN in depressive states, whereas imbalanced towards the SMN in manic states.^43,46^ While this remains to be tested, evidence of hyperconnectivity between the SMN and the DMN in bipolar patients in a depressed state seems to contradict this idea.^44^ As the balanced coupling between task positive and task negative networks is important for the proper execution of motor tasks in HC,^47^ we explore here whether the relationship between SMN-DMN networks can better explain possible FTT differences between groups.

Thus, in the present investigation, we expect that FTT motor performance is impaired in BD and UD in relation to HC, a difference that could decrease after antidepressant treatment. We hypothesize that the functional connectivity of M1 and PCC (our regions of interest for the seed analysis) with the SMA relates to FTT motor performance. Finally, we explore if the cross-correlation between the entire SMN and DMN (rather than the regions of interest) better explains differences in the level of motor performance.

## 2. Materials and methods

### 2.1 Protocol

The study was performed at the University Medical Center Göttingen (UMG) after approval of the research protocol by the local Ethics Committee. All participants provided their verbal and written informed consent after a detailed explanation of the research protocol. Patients experiencing a depressive phase of major depressive disorder (UD) or bipolar disorder (BD) (cf. ICD-10) and sex, age and education matched HC were recruited by announcements at the local University, the UMG Psychiatric Center as well as the local Asklepios Psychiatric Hospital.

All participants were between 18 and 65 years old. Exclusion criteria were: current or previous brain injury, current or past neurological disease, current use of illegal drugs, inability to perform the tasks or to undergo MRI sessions.

We collected individual information about the handedness^48^ and being lefthanded was not an exclusion criterion in order to consider all the handedness variability in the clinical sample. And to consider the influence of this variable, we also complement the analyses with models excluding lefthanders.

The study comprised two assessments of two consecutive days separated by approximately 5 weeks (Fig. 1). Between evaluations, patients received psychopharmacological treatment for the depressive episode at the psychiatric hospital by their caregiver physicians, who were independent in their treatment choices from the research group that conducted the study (naturalistic design). HC participated in the same study protocol, but received no treatment. All subjects were instructed to abstain from alcohol for at least 24 hours before and to refrain from consuming nicotine or caffeine for at least two hours before the assessments. On each day the participants performed the FTT, on the second day followed by a structured interview led by a trained person and – in case of medication with measurable blood metabolites (Supplementary Table 1) – a blood sample was taken immediately prior to the MRI session to measure their blood medication levels.

**Figure 1.**
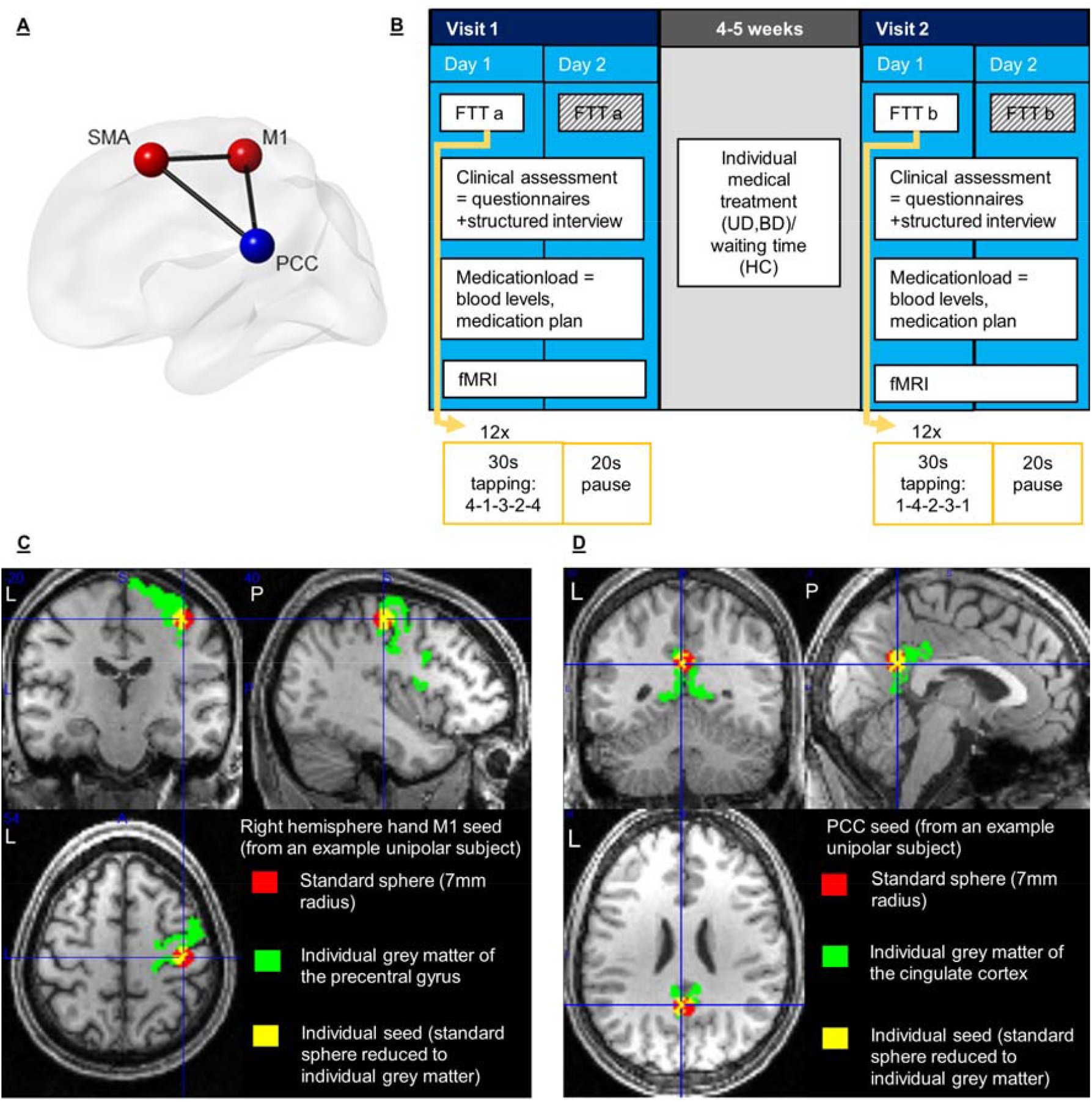
Methods. **(A)** Connectivity model between regions of interest: supplementary motor area (SMA) and right hemisphere hand primary motor cortex (M1) as part of the sensory motor network (red), posterior cingulate cortex (PCC) as part of the default mode network (blue); **(B)** Study design: unipolar (UD), bipolar (BD) depressed and healthy control subjects (HC) performed the finger-tapping-task (FTT) on both days of both visits, FTTa and FTT b versions as described in methods section. The data of the second days was not used for the analysis of motor performance. The clinical assessment and the MRI sessions took place on one of the days. The blood sample was taken as close as possible before the MRI measurement; **(C)** Seed definition technique: exemplification of the overlapping area (yellow) based on FreeSurfer individual segmentation of grey matter (green) and a spherical seed (red) at the MNI coordinates x = 40, y = -20, z = 54 of functional coordinates for right hemisphere hand motor cortex (M1)^59^ **(D)** and at x= 0 y = -53, z = 2 for the posterior cingulate cortex^17^.

Severity of symptoms in patients was assessed with the Montgomery Åsberg Depression Rating Scale (MADRS), the Young-Mania-Rating-Scale (YMRS), and the Beck-Depression-Rating-Scale second version (BDI-II), the first two rated within the structured interview, the latter, self-rated. We used the BDI-II in all our groups as our primary scale to implement the degree of depressive symptomatology in the analyses. The BDI-II is well correlated with other-rated scales (MADRS, Hamilton depression rating scale (HAMD)) and has a focus on cognitive aspects of depression and less on mood and anxiety or neurovegetative aspects,^49^ which is in line with the research focus of this study. Furthermore, the BDI-II as a self-rated scale showed more correlation to brain metabolism in resting state positron emission tomography than HAMD as an other-rated scale, indicating that BDI-II can be more strongly related to basal brain functioning.^50^ To account for the amount of psychotropic medication used per patient, an extended concept previously established as medication load^51–55^ was implemented (Supplementary material, chapter 1.1). In HC, to verify the absence of potential psychiatric symptoms, the self-rated Symptom-Checklist-90-R (SCL-90-R) was used.

### 2.2 Finger-Tapping-Task

The FTT was performed on a computer with an adapted keyboard that contained only the keys 1 to 4. The subjects tapped with the digits of the non-dominant hand not using the thumb. The task consisted of 12 intervals of 30 seconds duration in which the tapping was performed with intervals of 20 seconds without digit movements between them. The entire tapping sequence was presented on the computer monitor with numbers associated with digits: 1 = little finger, 2 = ring finger, 3 = middle finger, 4 = index finger. On the two days of the first visit the sequence “4-1-3-2-4” was used, on the two days of the second visit the sequence “1-4-2-3-1” was used (Fig. 1). The participants were instructed to press the keys as fast and accurately as possible. No feedback was given on the performance of the task. The mean from the number of correctly tapped sequences of the last three of the 12 tapping intervals of the first days of the visits where used as a score to quantify the motor performance – as they are expected to represent the performance plateau.^36,56^ The number of correctly typed sequences accounts for speed and accuracy.^35,36^

### 2.3 Functional connectivity

The functional and structural images were acquired with a 3T Siemens Magnetom Tim Trio, on Syngo VB-17a (Erlangen, Germany) and a 32-channel head coil. Structural whole-brain T1-weighted images were realized with a magnetization prepared rapid gradient echo (MPRAGE) sequence (repetition time: 2250 ms, echo time: 3.26 ms, inversion time: 900 ms, flip angle: 9°, field or view: 256 mm, spatial resolution: 1 mm x 1 mm, 176 slices, acquisition time: 8:26 min) and a resulting voxel size of 1mm x1mm x1 mm.

Resting state functional MRI (rs-fMRI) was measured with an gradient echo EPI (echo planar imaging) sequence (repetition time: 2500 ms, echo time: 33 ms, flip angle 70°, multi-band-factor 3, field of view 210 mm, spatial resolution: 2 mm x 2 mm, 60 slides with 2 mm thickness, distance factor 10%, 125 volumes (first 5 volumes deleted)) with a total acquisition time of 5:25 min, voxel size of 2 mm x 2 mm x 2 mm and a slice orientation along the anterior-posterior commissure.

Brain images were analyzed with SPM12 (www.fil.ion.ucl.ac.uk/spm, Wellcome Trust Centre for Neuroimaging, London, United Kingdom) based on MATLAB 2015b (MathWorks, Natick, Massachusetts) The following preprocessing steps were performed: default temporal high-pass filtering SPM12 at <0.008 Hz, slice-time-correction, realignment with unwarping SPM12; spatial normalization with the deformation field from FreeSurfer Version 6.0 (https://surfer.nmr.mgh.harvard.edu/) run on an Ubuntu 18.0 to the standard Montreal Neurological Institute (MNI) 152 brain at 2 mm spatial resolution; spatial smoothing with a Gaussian kernel of 6 mm full width at half maximum (FWHM); nuisance-regression of white matter and cerebral spinal fluid as well as motion correction with an automatized independent component analysis based cleaning procedure (ICA-AROMA v0.3-bet, available from: https://github.com/maartenmennes/ICA-AROMA).^57^ To ensure all autocorrelative signal was regressed out, a visual inspection of all data for quality control was performed. If potential noise artefacts remained, they were added as additional “noise” components to the automatically identified by ICA-AROMA and the cleaning procedure was rerun. In addition to this extensive correction of movement we also assessed potential bias due to different extend of movement between the groups by comparing individual root mean squares based on frame-by-frame displacement of adjacent volumes in mm between the groups in an repeated measures ANOVA (rmANOVA).^58^

Seed-based analyses aimed to address functional connectivity differences and its influences on motor performance of the right hemisphere hand M1 and the PCC, especially to the SMA as the core region of our hypotheses. To define the M1 gray matter seeds, we combined the anatomical T1 information from the precentral gyrus from FreeSurfer segmentation with a 7 mm radius spherical seed, defined using the SPM toolbox MarsBaR version 0.44 (http://marsbar.sourceforge.net/), which was centered at the MNI-coordinates 40 -20 54 (x, y, z) for the right hemisphere hand M1 from a meta-analysis of functional MRI data of hand movement tasks.^59^ The overlap between these two sources was carefully inspected for consistency in every individual subject’s data (see an example for precise coverage of the grey matter with a seed based on this technique in Fig. 1). To make sure that differences in functional connectivity did not stem from differences in the seed volume, we checked if the volume of the individualized masks differed between groups or time with an rmANOVA in SPSS (for results see Supplementary material, chapter 2.2). The newly established seed definition technique was also applied to create seeds for the PCC (MNI coordinates 0 -53 26 (x, y, z)) (for an example see Fig. 1), centered in the position used for the investigation of motor impairments in stroke.^17^

The time courses of these seeds were bandpass filtered (0.01 Hz – 0.1 Hz) before being included into a general linear model (GLM) in SPM12 for the first level analysis. According to the known task-positive and task-negative activations in the right hemisphere hand M1 and PCC respectively, we focused on brain regions positively correlated when seeding in the right hemisphere hand M1 and regions negatively correlated (anticorrelated) when seeding in the PCC.

To investigate potential full network interconnection deficiency between SMN and DMN in depressive patients, a group independent component analysis (gICA) was performed using MELODIC Version 3.14 in FSL (FMRIB’s Software Library, www.fmrib.ox.ac.uk/fsl),^60^ to be able to estimate correlation between the time courses of these networks in all groups before and after intervention. The gICAs were performed for each group and each visit separately. Then, all the components were inspected selecting the components reflecting SMN, anterior DMN and posterior DMN. The time course of the network signal from every subject was extracted separately for each selected component. For every subject a correlation coefficient for the correlation between the time courses of the SMN and the anterior DMN as well as for the correlation between the SMN and the posterior DMN was calculated using MATLAB 2015b (MathWorks, Natick, Massachusetts).

### 2.4 Statistical Analysis

The FTT was analyzed with an rmANOVA for motor performance in IBM SPSS Statistics Version 26. In the main analysis no covariates were included, but to check for influencing effects, the analysis was repeated with the covariates sex, age, education, change of the medication load and change in BDI-II, which have the potential to influence the motor performance.^25,61–65^ Furthermore, models with only the patient groups were created to consider potential effects of years since first episode, age of onset, number of depressive episodes, change in MADRS and change in YMRS, which have been described in the literature.^29,63^ The statistical threshold for the tests was set to two-tailed *p < 0.05*. If the assumption of homogeneity of variance was met for the ANOVAs, we used the Bonferroni post-hoc-test to correct for multiple testing. But if this assumptions was not met, we used the Games-Howell post-hoc-test.

For the seed based rs-fMRI data, two SPM full factorial models with group and time as factors were built for the two seeds in the second level analysis, one without covariates and one with medication load as covariate-of-no-interest to control for potential differences stemming from the load of medication the patients received. In addition, linear regressions with functional connectivity of the seed and motor performance were calculated for the whole study population.

The correlation coefficients were used for an rmANOVA model calculated in SPSS to investigate whether possible correlations between the networks differed between groups or time. Additionally, a linear regression was performed for each visit to test for a relationship between the SMN-DMN time course correlations and the motor performance.

In regions belonging to our a priori hypothesis, we considered the statistical threshold at the cluster-wise correction level (*p FWEc <0.05*) to reduce the probability of both false-positive and false-negative results. Otherwise, we considered the family-wise error correction level *(p FWE < 0.05*) to reduce the probability of false-positive results. The labeling of functional imaging clusters was performed according to the automated anatomical labeling (AAL) atlas.^66^

### 2.5 Data availability

The data supporting the findings of this study are available from the corresponding author upon request. Due to restrictions in the data sharing consent obtained from the study participants the data are not publicly available.

## 3. Results

From 95 participants, 11 were unable or unwilling to complete the entire study, one participant performed the FTT with the wrong hand and four participants were excluded due to abnormalities in the anatomical scans, e.g. intracranial epidermoid cysts. Therefore, data from 79 participants (31 HC, 27 UD, 21 BD - 18 Type I, three Type II) were taken into final analysis. One participant from each of the HC and BD groups and two participants of the UD group were lefthanded.

### 3.1 Characterization of groups

The mean time between the two visits was 4.9 weeks (SD 0.8). A complete evaluation of the characteristics between groups is presented in Table 1. In line with our matching, the groups were comparable in terms of sex, age and years of education. The patient groups did not differ in terms of the history of their disease measured by age of onset, years since first episode and number of depressive episodes. The healthy controls did not exceed the threshold of normality in the global severity index of the SCL-90-R score.

**Table 1:** Characteristics of the bipolar (BD), unipolar (UD) and healthy control (HC) groups.

|  | *Controls* | *Unipolar* | *Bipolar* | *Comparison p-value* | *Meaning of p* |
| --- | --- | --- | --- | --- | --- |
| N (% female) | 31 (58.1%) | 27 (40.7%) | 21 (33.3%) | 0.176<sup>a</sup> | No difference between groups |
| Age in years | 41.7 ± 14.3 | 39.6 ± 15.3 | 43.5 ± 9.8 | 0.591<sup>b</sup> | No difference between groups |
| Education in years | 16.5 ± 3.8 | 15.5 ± 3.1 | 16.8 ± 4.0 | 0.453<sup>c</sup> | No difference between groups |
| Number lefthanders | 1 (3.2%) | 2 (7.4%) | 1 (4.8%) | X<sup>d</sup> | X<sup>d</sup> |
| Age of first episode | X | 29.5 ± 15.2 | 24.7 ± 9.5 | 0.182<sup>b</sup> | No difference between groups |
| Years since first episode | X | 10.1 ± 9.4 | 18.9 ± 13.5 | 0.161<sup>b</sup> | No difference between groups |
| Number of depressive episodes | X | 4.8 ± 3.8 | 7.2 ± 6.1 | 0.095<sup>c</sup> | No difference between groups |
| Number of manic episodes | X | X | 2.9 ± 3.6 | X | X |
| Number of hypomanic episodes | X | X | 3.3 ± 5.0 | X | X |
| BDI-II visit 1 | 2.6 ± 3.7 | 29.5 ± 11.1 | 21.1 ± 16.7 | <0.001<sup>b</sup> | Games-Howell-Post-Hoc-Test: Difference HC and UD: **p<0.001**, HC and BD: **p<0.001**, UD and BD: **p = 0.128** |
| BDI-II visit 2 | 2.3 ± 4.2 | 20.2 ± 14.0 | 17.1 ± 13.1 | <0.001<sup>b</sup> | Games-Howell-Post-Hoc-Test: Difference HC and UD: **p<0.001**, HC and BD: **p<0.001**, UD and BD: **p = 0.714** |
| Delta BDI-II visit 2 – visit 1 | -0.2 ± 2.6 | -9.3 ± 11.9 | -3.9 ± 13.3 | 0.009<sup>b</sup> | Games-Howell-Post-Hoc-Test: Difference HC and UD: **p = 0.002**, HC and BD: **p = 0.437**, UD and BD: **p = 0.327** |
| MADRS visit 1 | X | 24.2 ± 8.6 | 13.9 ± 10.6 | 0.001<sup>c</sup> | Severity of depressive symptoms is higher in UD |
| MADRS visit 2 | X | 14.3 ± 10.2 | 10.5 ± 8.3 | 0.179<sup>c</sup> | No difference between groups |
| Delta MADRS visit 2 – visit 1 | X | -9.9 ± 10.3 | -3.3 ± 9.3 | 0.028<sup>c</sup> | Reduction in the severity of depressive symptoms is higher in UD |
| YMRS visit 1 | X | 1.7 ± 2.6 | 6.1 ± 6.9 | 0.012<sup>b</sup> | Severity of manic symptoms (mixed episode) is higher in BD |
| YMRS visit 2 | X | 1.4 ± 1.7 | 2.3 ± 3.5 | 0.222<sup>b</sup> | No difference between groups |
| **Delta YMRS visit 2 – visit 1** | X | -0.3 ± 2.1 | -3.7 ± 6.2 | **0.025<sup>b</sup>** | Reduction in the severity of manic symptoms (mixed episode) is higher in BD |
| **Medication load visit 1** | 0 ± 0 | 4.3 ± 3 | 4.5 ± 2.4 | No p-value because of a variance of 0 in controls<sup>b</sup> | Games-Howell-Post-Hoc-Test: Difference HC and UD: **p<0.001**, HC and BD: **p<0.001**, UD and BD: p = 0.938 |
| **Medication load visit 2** | 0 ± 0 | 4.1 ± 2.6 | 3.8 ± 2.3 | No p-value because of a variance of 0 in controls<sup>b</sup> | Games-Howell-Post-Hoc-Test: Difference HC and UD: **p<0.001**, HC and BD: **p<0.001**, UD and BD: p = 0.925 |
| **Delta medication load visit 2 – visit 1** | 0 ± 0 | -0.2 ± 1.9 | -0.7 ± 2.5 | No p-value because of a variance of 0 in controls<sup>b</sup> | Games-Howell-Post-Hoc-Test: Difference HC and UD: p = 0.867, HC and BD: p = 0.405, UD and BD: p = 0.700 |

<sup>a</sup>Chi-Square-Test-Pearson
<sup>b</sup>One-way-ANOVA with Brown-Forsythe correction for inhomogeneity of variance
<sup>c</sup>One-way-ANOVA with homogeneity of variance
<sup>d</sup>Analysis repeated without lefthanders
Abbreviations: BD = Bipolar depressed; BDI-II = Beck-Depression-Rating-Scale second version; HC = Healthy controls; MADRS = Montgomery Åsberg Depression Rating Scale; UD = Unipolar depressed ; YMRS = Young-Mania-Rating-Scale

The severity of the depressive episode was evaluated during the study and at both visits the patient groups showed comparable depressive episode severity according to the BDI-II scores. The comparison of the difference in BDI-II scores of visit 1 and 2 between groups revealed a significantly higher difference only for UD compared to HC (Table 1). An rmANOVA with the BDI-II scores revealed a significant change between visit 1 and 2 that seems driven by a stronger reduction in the patient groups (factor “time *F(1,76) = 15.660, p < 0.001*; interaction “group x time” *F(2,76) = 6.035, p = 0.004*; factor “group” *F(2,76) = 41.138, p < 0.001*).

In MADRS, the severity of depressive symptoms was significantly higher in UD, but only at the first visit (Table 1). An rmANOVA with the MADRS scores revealed a significant reduction between visit 1 and 2 for both patient groups but stronger in UD than in BD (factor “time” *F(1,46) = 21.067, p < 0.001*; interaction “group x time” *F(1,46) = 5.179, p = 0.028*; factor “group” *F(1,46) = 8.903, p = 0.005*). There was a significant difference seen for YMRS scores with BD showing higher scores and a stronger reduction across time than UD in an rmANOVA (factor “time *F(1,46) = 10.067, p = 0.003*; interaction “group x time” *F(1,46) = 7.024, p = 0.011*; factor “group” *F(1,46) = 7.491, p = 0.009*).

Although only bipolar patients in the current depressive episode were recruited, the symptoms were mixed in some patients. At the first visit, three bipolar patients showed subclinical depressive symptoms (*BDI-II score* ≤ *8* and *MADRS score* ≤ *12*), five showed manic symptoms (*YMRS score > 9*) and one showed both manic (*YMRS score of this subject at visit 1 = 14*) and depressive (*BDI-II score of this subject at visit 1 = 53*; *MADRS score of this subject at visit 1 = 28*) symptoms. Even though being subclinical (*M*_*YMRS*_ ≤ *9*), the BD group showed a significantly higher YMRS score on visit 1 compared to the UD, as well as a greater change in YMRS score between the two visits. To confirm that the main results were not driven by the individuals with manic symptoms characteristics or subclinical depressive symptoms characteristics, we compared mean motor performance scores of the original BD group, with a subgroup of BD with clinical depressive symptoms only, which showed comparable values (*BD visit 1 = 8.6 ± 4.6 (M ± SD*) (Fig. 2); BD visit 1 with clinical depressive symptoms only: *8.4 ± 4.9 (M ± SD)*). We also repeated the imaging analysis of linear regression of motor performance scores and functional connectivity of PCC seeds without the BD subjects showing manic, mixed or subclinical symptoms and the main results could still be identified.

**Figure 2.**
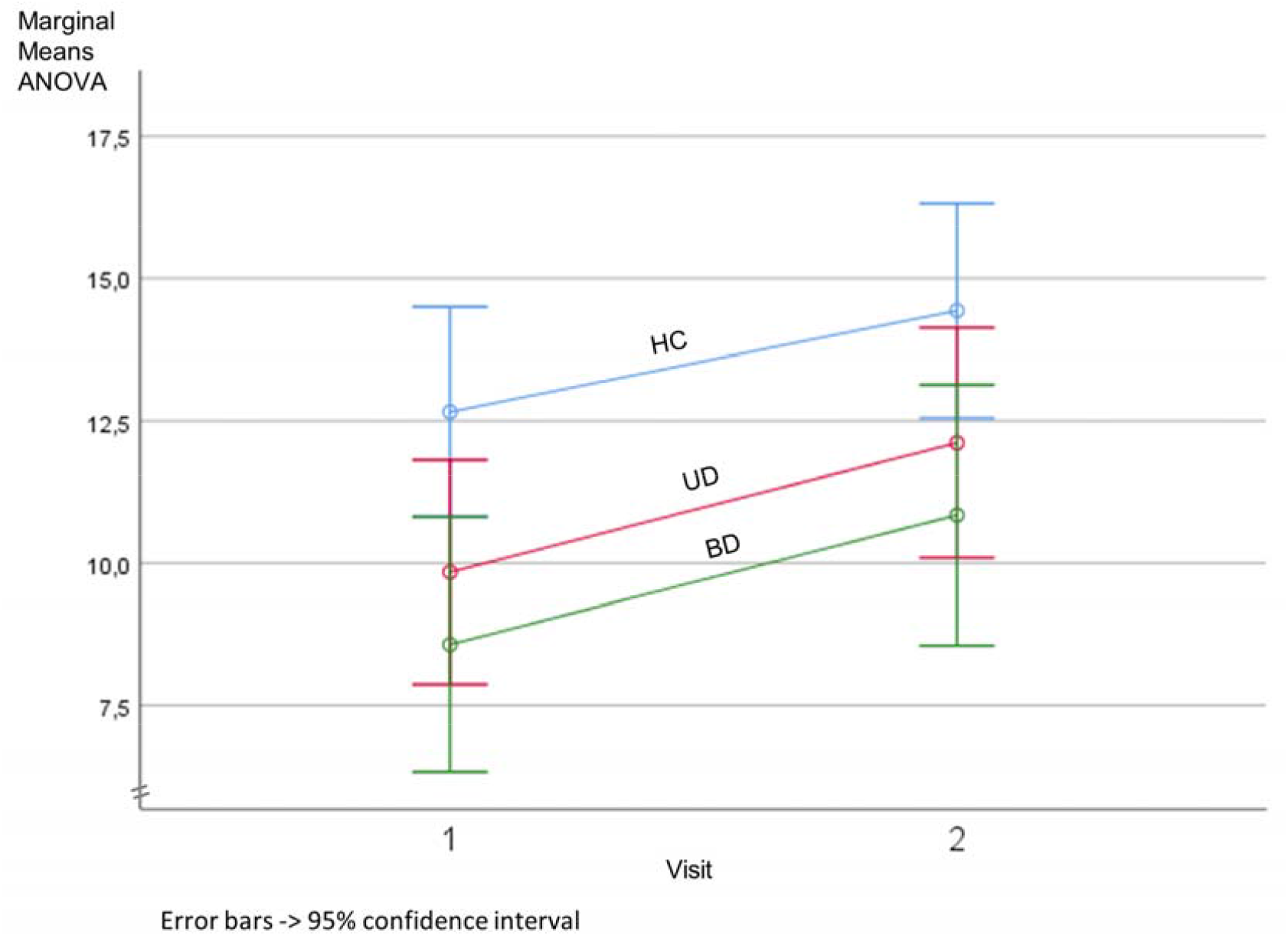
Motor performance scores. Marginal means (mean number of correctly tapped sequences per 30 seconds) and confidence intervals from repeated-measures ANOVA of motor performance scores for healthy controls (HC, blue), unipolar (UD, pink) and bipolar (BD, green) patients. The values displayed in the graph are HC visit 1 *M ± SD = 12.7 ± 6.1*, UD visit 1 *M ± SD = 9.8 ± 4.2*, BD visit 1 *M ± SD = 8.6 ± 4.6*, HC visit 2 *M ± SD = 14.4 ± 5.8*, UD visit 2 *M ± SD = 12.1 ± 5.0*, BD visit 2 *M ± SD = 10.8 ± 4.7*. Significant differences are seen only between HC and BD (ANOVA factor “group” *F(2,76) = 4.122*; *p = 0.020*) with BD showing a significantly reduced number of correctly performed finger tapping trials compared to the HC (Post-hoc-test *p = 0.023*).

No difference in medication load between patient groups was seen at visit 1 or visit 2, but the load in both visits was significantly higher compared to HC, since they were not medicated. Furthermore, the change in medication load between visit 1 and 2 was small in the patient groups (Table 1) and not distinguishable from the HC on an additional rmANOVA (factor “time” *F(1,76) = 2.431, p = 0.123*; interaction “group x time” *F(2,76) = 1.155, p = 0.320*; factor “group” *F(2,76) = 48.226, p < 0.001*). For a detailed description of medication taken by the patient groups see tables of chapter 2.1 in the Supplementary material.

### 3.2 Behavioral results

The motor performance differed between groups (ANOVA factor “group” *F(2,76) = 4.122*; *p = 0.020*; Fig. 2) with BD showing a significantly reduced number of correctly performed finger tapping trials compared to the HC (Post-hoc-test *p = 0.023*). The UD group showed no performance impairment compared to HC (Post-hoc-test *p = 0.158*). The BD and UD showed no significant difference (Post-hoc-test *p = 1*).

All groups showed an improvement of their motor performance in visit 2 (ANOVA factor “time” *F(1,76) = 33.800*; *p < 0.001*). The improvement did not differ between the groups (interaction “group x time” *F(2,76) = 0.230*; *p = 0.795*).

By including the covariates sex, age, education, change of medication load and change in BDI-II into the model, the difference in performance between the groups remains significantly different (factor “group” *F(2,71) = 5.194*; *p = 0.008*) as does the comparable performance improvement of the groups across the two visits (interaction “group x time” *F(2,71) = 0.067*; *p = 0.935*). However, the general effect of the performance improvement seen across the groups in visit 2 was not present anymore (factor “time” *F(1,71) = 0.002*; *p = 0.966*).

Influences of covariates on the motor performance in general revealed a negative correlation with age (*F(1,71) = 23.542*; *p < 0.001*) and a positive correlation with education (*F(1,71) = 16.183*; *p < 0.001*) for all groups. When limiting the ANOVA to the patient groups, an improvement across visits was present (factor “time” *F(1,46) = 23.042*; *p < 0.001*), which did not differ between the groups (factor “group”; *F(1,46) = 1.010*; *p = 0.320*) nor showed an interaction with visits (interaction “group” x “time”, *F(1,46) < 0.001*; *p = 0.999*). These results were maintained, when including the covariates years since first episode, age of onset, number of depressive episodes, change in MADRS and change in YMRS. An older age of first episode (*F(1,41) = 5.808*; *p = 0.021*) and more years since first episode (*F(1,41) = 4.396*; *p = 0.042*) reduced the motor performance. An increase in YMRS score was positively correlated with a change in motor performance (interaction time x deltaYMRS *F(1,41) = 4.525*; *p = 0.039*), a change in MADRS was not (interaction “time x deltaMADRS” *F(1,41) = 0.799*; *p = 0.377*). The number of depressive episodes showed a negative correlation with change in performance (interaction “time x number of depressive episodes” *F(1,41) = 8.489*; *p = 0.006*).

All results hold, when excluding the lefthanders from the analysis.

### 3.3 Functional connectivity

#### 3.3.1 PCC seed functional connectivity

For each group, the seed based functional connectivity between the PCC and the SMA was visible as a negative correlation at *p FWE < 0.05* level, except for the BD group at visit 2. This is in contrast to the seed based connectivity between PCC and M1, were no negative correlation was identified at *p FWE < 0.05* level in all groups and visits. Positive correlation to the PCC was evidenced for regions of the DMN, namely angular gyri, precuneus, cingulate gyri, and middle prefrontal regions (data not shown).

The connectivity between PCC and SMA was lower at visit 2 for BD in contrast to HC, as well as to UD, at *p FWEc < 0.05* level (Fig. 3). This remains the same, when excluding the lefthanders from the model. No group differences in the functional PCC-SMA connectivity were observed at visit 1. In addition, no differences in connectivity of PCC to M1 were observed between groups or time. Lastly, all findings from the PCC seed analysis are sustained, when medication load is included in the full factorial models as a regressor-of-no-interest.

**Figure 3.**
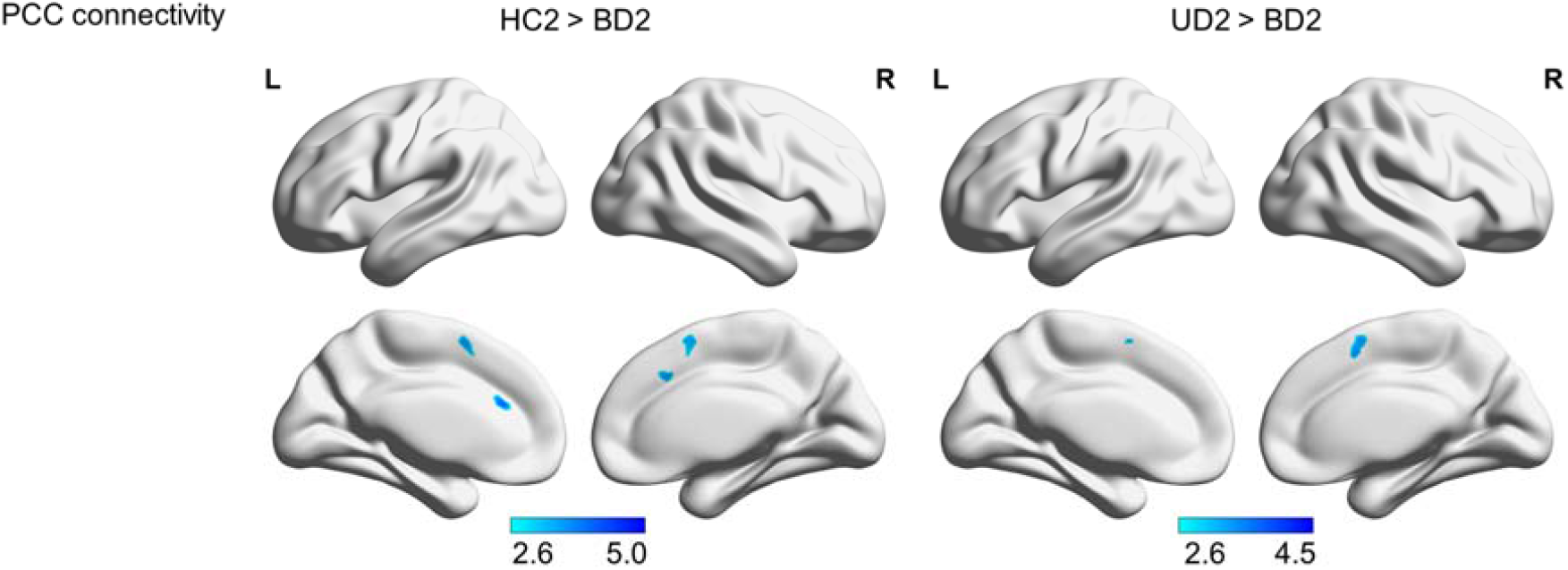
PCC connectivity differences. Resting state functional connectivity differences of the posterior cingulate cortex (PCC) at the second visit between the bipolar depressive subjects (BD2) and the unipolar depressive (UD2) resp. control (HC2) subjects (p FWEc < 0.05). HC2 showed higher connectivity measured by anticorrelation between the PCC and the bilateral SMA than BD2. UD2 showed higher connectivity measured by anticorrelation between the PCC and the bilateral SMA than BD2. Colorbar represents t-values.

The linear regression of the PCC-SMA functional connectivity and the motor performance scores revealed a positive correlation, i.e. the higher the performance the stronger the negative functional connectivity between PCC-SMA (Fig.4). When excluding the lefthanders from analysis the cluster in the SMA maintained at a level of *p <0.001* uncorrected, but did not survive *FWEc < 0.05* level anymore, most likely due to the reduced power by the lower number of subjects in the analysis.

On the other hand, the linear regression of the PCC-M1 functional connectivity and the motor performance scores showed no linear relationship. Above all, supporting the idea that lower PCC-SMA anticorrelation in BD is related to their behavioral impairment, a partial overlap in SMA is seen between the functional connectivity differences with other groups and the linear regression with motor performance (Fig. 5). In other words, part of the SMA in BD relates to both reduced performance in FTT and reduced functional connectivity to PCC, not M1.

**Figure 4.**
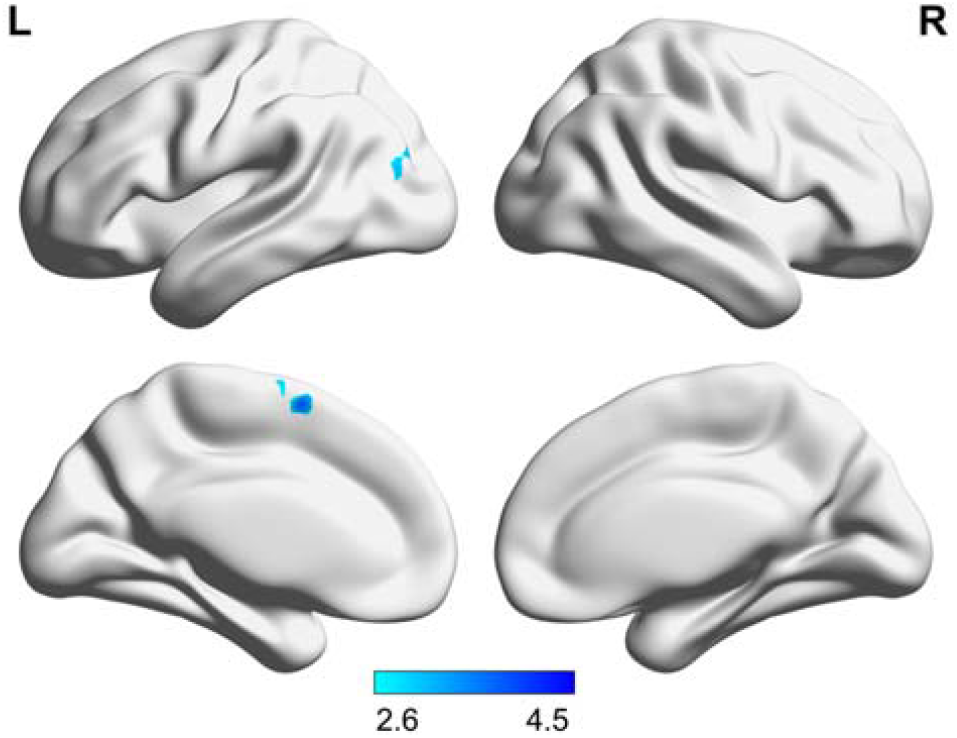
PCC connectivity correlated with motor performance. Resting state functional connectivity of the posterior cingulate cortex (PCC) linearly correlated with the motor performance in the finger-tapping-task (p FWEc < 0.05). The motor performance is correlated with the connectivity (anticorrelation) strength between the PCC and the left supplementary motor area (SMA). Colorbar represents t-values.

**Figure 5.**
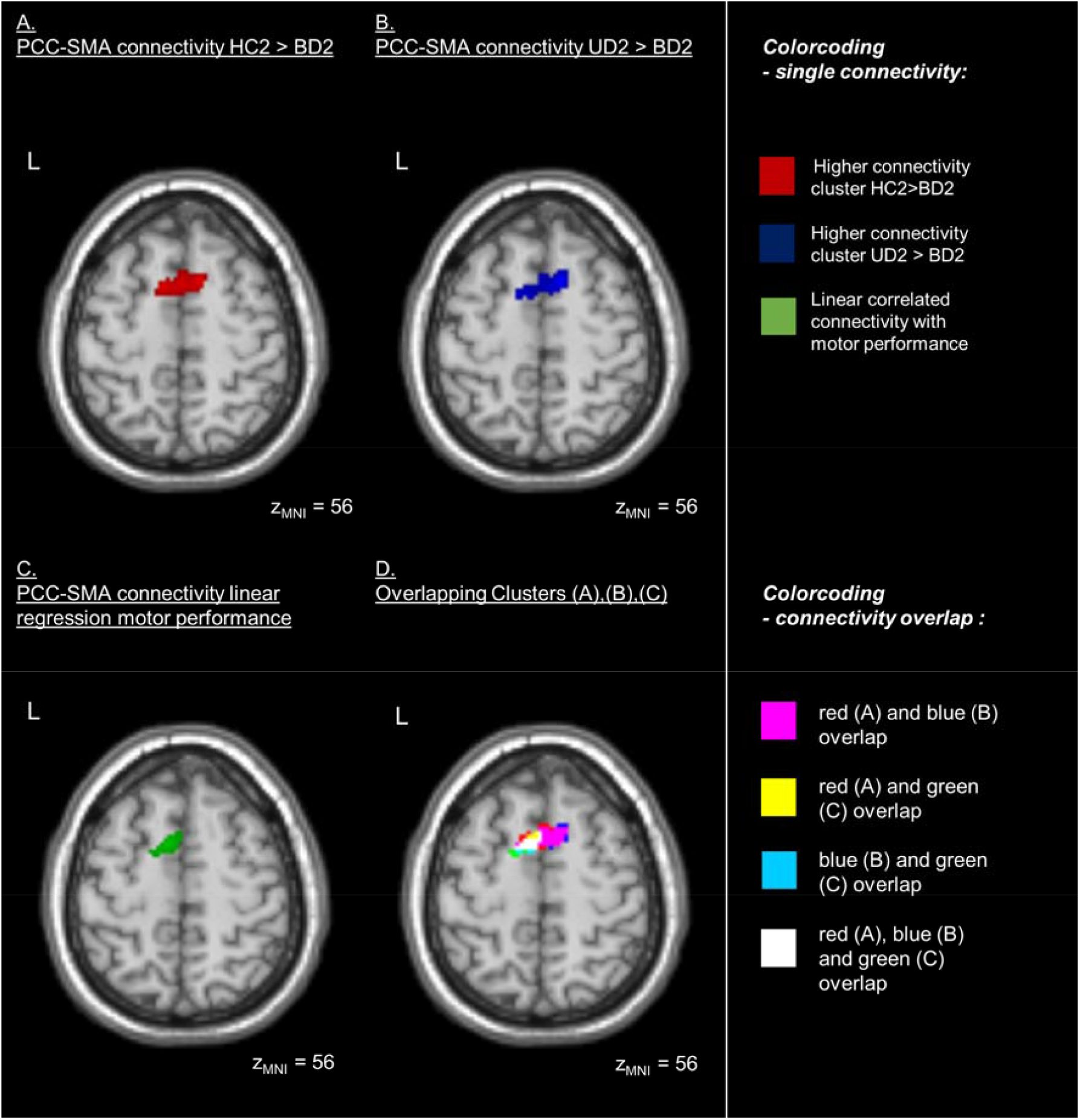
PCC connectivity overlaps in the SMA. Resting state functional connectivity differences of the posterior cingulate cortex (PCC) between groups and connectivity of the PCC correlated with motor performance overlap in the supplementary motor area (SMA) at an axial plane (z_MNI_ = 56, *p FWEc < 0.05*). **(A)** Healthy control subjects (HC2) show higher connectivity than the bipolar depressive subjects (BD2) at visit 2. **(B)** Unipolar depressive subjects (UD2) present higher connectivity than the bipolar depressive subjects (BD2) at visit 2. **(C)** The connectivity of the PCC to a part of the left SMA is correlated with the motor performance in the finger-tapping-task. **(D)** The SMA connectivity clusters seen in (A, red), (B, blue) and (C, green) overlap in different extensions, as represented in pink (A⋂B), yellow (A⋂C), turquoise (B⋂C) and white (A⋂B⋂C).

#### 3.3.1 M1 seed based functional connectivity

The positive functional connectivity between the right hemisphere hand M1 and the SMA could be shown at *p FWE < 0.05* for each of the groups. Additionally, other parts of the SMN, namely the pre- and postcentral gyrus bilaterally as well as bilateral temporal regions were identified. No negative correlation was evidenced at *p FWE < 0.05* (data not shown).

A higher connectivity between the M1 and the SMA was seen in HC compared to UD only at visit 2 (*p FWEc < 0.05*) in the model where the medication load was inserted as regressor-of-no-interest. When excluding the lefthanders from the model with medication load as a regressor-of-no-interest, a higher connectivity between the right hemisphere hand M1 and the SMA was also seen at visit 1 at *p FWEc< 0.05* threshold.

Compared to UD, BD showed stronger connectivity between M1 and SMA on both visits (Fig. 6). At the first visit, these clusters survived *p FWE < 0.05* correction. At the second visit, differences were more restricted and seen only in the left hemisphere at *p FWEc < 0.05* level. These differences between BD and UD held in the model with medication load as a regressor-of-no-interest as well as in the model without lefthanders. There was no connectivity of the M1 that was stronger in UD than in BD.

**Figure 6.**
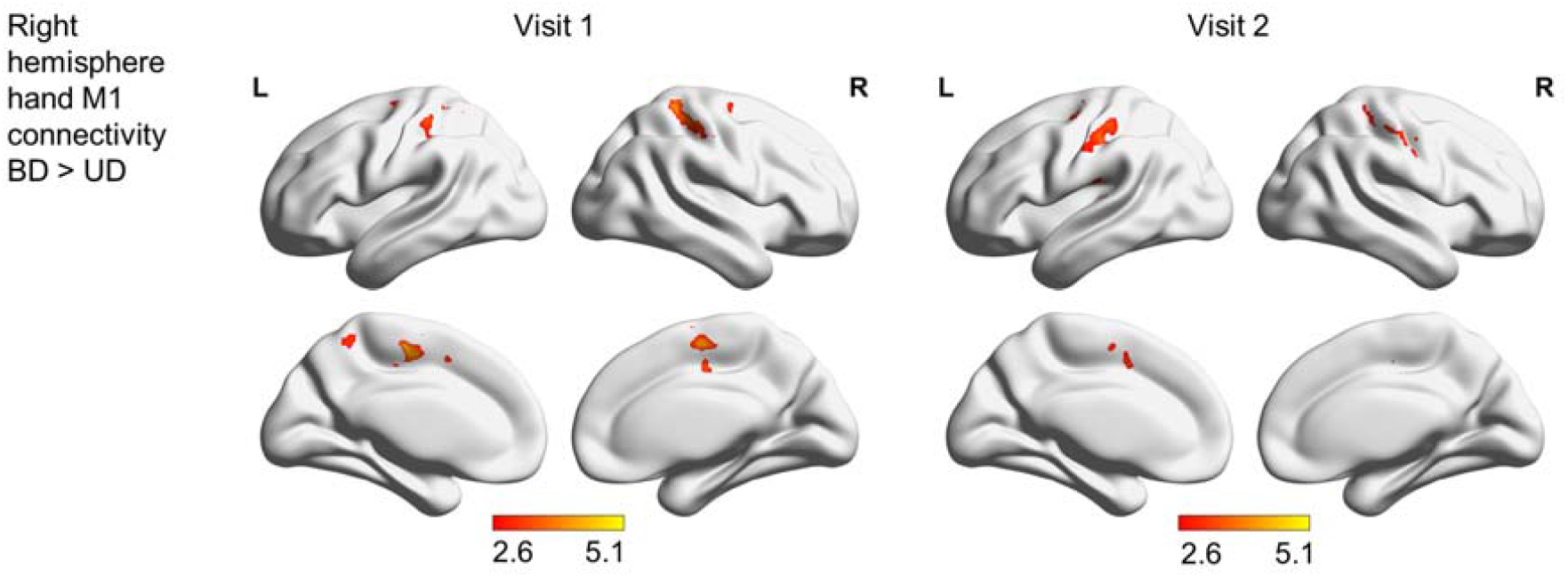
M1 connectivity difference. Differences in the resting state functional connectivity of the right hemisphere hand motor area (M1) between bipolar (BD) and unipolar (UD) depressed participants at *p FWEc < 0.05*. BD showed stronger connectivity than UD between M1 and the bilateral SMA in visit 1 than in visit 2. No regions were evidenced in the UD>BD contrast. Colorbar represents t-values.

The linear regression of motor performance score and the right hemisphere hand M1-SMA functional connectivity did not show SMA clusters that survived *p FWEc <0.05* correction, i.e. there was no correlation between the motor performance and functional connectivity between SMA and M1. This result remained when excluding the lefthanders from the model. All groups did not show changes of their right hemisphere hand M1 connectivity across time.

Beyond our a priori hypotheses, higher connectivity in HC than UD at visit 2 between the right hemisphere hand M1 and the right Rolandic operculum was seen (p FWE < 0.05). When controlling for the medication load as a regressor-of-no-interest, the cluster at the right Rolandic operculum was not seen. Furthermore, the cluster was not present, when excluding the lefthanders from the analysis. There was no further difference between BD and HC in any of these models.

Finally, to check for potential bias in the seed model, we evaluated whether differences in movement exists during the scanning sessions, in spite of applying a strict motion correction procedure in the analysis. When comparing the frame-by-frame displacement with root-mean-squares ^58^ between the three groups, a difference was revealed, which do not seem to stem from a difference between the HC-BD groups (post-hoc *p = 0.257*), but rather from a trend between the UD-BD groups (post-hoc *p = 0.062*) (for detailed results see Supplementary material, chapter 2.3).

### 3.4 SMN-DMN correlation coefficients

The correlation coefficients between SMN and DMN were computed and compared between groups and across time in rmANOVAs (for results see Supplementary material, chapter 2.4, Table 6). In the linear regressions, the motor performance scores neither showed a relationship with the correlation of SMN and posterior DMN (visit 1: *r < 0.001, p = 0.903*; visit 2: *r = 0.001, p = 0.772*), nor with the correlation of SMN and anterior DMN on any of the visits (visit 1: *r = 0.005, p = 0.375*; visit 2: *r = 0.002, p = 0.700*). Similar to the models with lefthanders, no linear relationship between SMN-DMN correlation and motor performance was seen for visit 1 or visit 2.

## 4. Discussion

The current study examined the neural underpinnings of motor performance between groups of BD, UD and HC participants with comparable sex, age, education, medication load and history of disease. We identified a deficit in the FFT motor performance only in BD in relation to HC, which decreased after five weeks of psychopharmacotherapy. As expected, an association between the FTT motor performance and the negative functional connectivity between PCC and SMA was evidenced, which in the explorative investigation did not extend to the relationship between DMN-SMN correlations. Contrary to our expectations, an association between FTT motor performance and positive functional connectivity between M1 and SMA was not observed, suggesting that the contribution of M1 is less relevant than that of PCC in this case.

Although the motor circuit is one of the best described in the literature,^for review see 67^ functional brain alterations in motor areas as part of depressive episodes remain poorly explored. In line with a former study showing the importance of the PCC-SMA connectivity for upper limb motor tasks,^17^ we were able to show here that this connectivity also appears critical for the FTT performance. Since connectivity between PCC-SMA is linearly correlated with the FTT performance, which is impaired in BD compared to HC, one can assume that the functional brain difference between groups would explain the behavioral difference in the FTT. Despite these promising findings, one must consider that such results do not survive multiple testing correction (*FWEc<0.05*), when excluding the lefthanders, although they remain identifiable at the threshold *p<0.001*. Rather than the lack of such connectivity effects in the model without lefthanders, the existence of these findings at this threshold supports the notion of a drop in power in detecting statistical differences. To our knowledge, this is the first time that PCC-SMA connectivity has been evaluated and identified as critical for motor performance in a study involving depressive and healthy subjects.

Psychomotor alterations are commonly seen in BD and sometimes in UD compared to HC, which has been suggested as a factor that negatively contributes to treatment response or even remission.^24,25^ Therefore, this remains a very relevant topic of research. In line with a former study comparing psychomotor performance in UD, BD and HC,^33^ we report evidence that BD are more likely to have an impairment in motor performance measured by the FTT. This finding is also in line with the literature showing more robust FTT impairment in BD^18,19^ than in UD, where it appears compromised only in some study samples.^5,20,21,37–39^ Overall, learning effects seem to be present from the first FTT assessment to the second, differently from previously shown.^68^ In a keyboard naive population from Kenya, it was seen that such learning effects only appeared in healthy controls and not in depressive subjects.^39^ A possible explanation for the improvement in the FTT performance could be that depressive patients in our study were able to profit from treatment, which may have reflected at least partially in their performance. This is an interesting hypothesis involving possible neuroplasticity changes that could be addressed in the future.

The hypothesis that the resting state functional connectivity between the M1 and the SMA is not only critical for the motor performance in HC,^42^ but also in UD and BD, was rejected since the linear regressions with motor performance did not show correlated clusters in the SMA when seeding the right hemisphere hand M1. As far as we know, this work is the first providing evidence that right hemisphere hand M1-SMA connectivity is not critical for influencing FTT motor performance in depressive subjects. Another possibility for negative findings is that the seed method in Herszage *et al*.^42^ when detecting the functional representation of the hand with transcranial magnetic stimulation for each subject, could have been more precise. Since our method of seed determination includes not only functional but also individual anatomical information, it would be interesting to compare the two seeding methods in future studies to define the most promising way of detecting individual seeds.

Our results show that BD have a higher M1-SMA connectivity than UD at both timepoints. Since we did not find behavioral differences in the FTT between the two patient groups, we can only hypothesize that the M1-SMA connectivity might have relevance for other symptoms of depression that were beyond the scope of this work. Groups, both BD and UD were comparably depressed, in a moderate level according to BDI-II scores, but only the UD improved significantly in the second evaluation, which might be a limitation when comparing the longitudinal changes during treatment. We speculate that residual psychomotor symptoms in BD, including the differences seen in neural correlates, may contribute for the limited improvement since psychomotor retardation is known as a negative predictor for antidepressant treatment response.^69^ All individual seeds used for M1 and PCC functional connectivity analysis have been carefully created and inspected for structural accuracy of each individual anatomy aiming at the participant’s hand knob. Furthermore, their volumes did not differ across groups, which per se could lead to false conclusions. Finally, we carefully controlled our main findings for medication load and movement during scanning sessions, where we found no evidence that our main findings might have been biased from these covariates. A trend difference of movement between groups should be kept in mind. To best remove potential bias stemming from movement, we used movement correction with ICA-AROMA which was shown to be very efficient^70^ and extended it with manual checking for artificial signals, so we believe that our results from BD-UD comparison are not biased by movement. In our view, our results also open new research perspectives for the comparisons between patient groups. For example, it may be interesting in the future to use motor paradigms that comprise a motor planning component, which may be more sensitive in detecting differences in behavior between UD and BD. This approach can be useful in new ways to support early differential diagnosis where there is a clear clinical need.^71^

It was postulated that the imbalance between the SMN and the DMN is critical for motor performance in depression.^for review see 6^ Therefore, it might be expected that the interconnection between the SMN and DMN, as a task positive and a task negative network respectively^47,72^, is critical for motor performance in the FTT. To explore whether this model could explain the motor performance differences in a more consistent way than our seed model would, we computed individual correlation coefficients between SMN and DMN. The idea that the SMN-DMN connectivity is critical for FTT was rejected, since no linear relationship between correlation coefficients and the motor performance were revealed. It is possible that a model taking into account the imbalance between whole networks containing many regions is likely to oversimplify more complex neural mechanisms in this case.

Some potential limitations need to be mentioned, which make our results be viewed with caution. Our sample size is comparable to other studies, but likely modest taking into account the limited power in detecting differences when excluding the lefthanders. Therefore, a replication of our findings with larger samples is prudent. Another important aspect is the use of medication, which has been very tightly controlled, but still could have influenced the results in unpredictable ways. Probably due to fast switches in mood, not all BD were rated with depressive symptoms at a clinical threshold at the day of testing and some also showed a combination of depressive and manic symptoms (mixed episode), which might be a limitation of this study but reflects clinical reality. Noteworthy, considering that BD can be often misdiagnosed as UD,^71^ we observed the occurrence of manic symptoms in one patient part of the UD group, for whom the YMRS was in the clinical range. Nevertheless, all patients included in this study had a long history of disease and a diagnosis confirmed by multiple caregivers of the clinical setting, which reduces the risk of a BD being misdiagnosed as UD.71,73

In conclusion, our seed model supports the notion that functional connectivity between PCC-SMA explains the FTT performance in healthy and depressed individuals, but the functional connectivity between M1-SMA does not. Reinforcing the relevance and originality of these findings, the SMN-DMN correlation did not explain motor performance as the seed model.

## Data Availability

All data produced in the present work are contained in the manuscript.

## Abbreviations

AAL: automated anatomical labeling
BD: Bipolar depressed
BDI-II: Beck-Depression-Rating-Scale second version
DMN: Default mode network
EPI: Echo planar imaging
FTT: Finger-tapping-task
gICA: group independent component analysis
GLM: general linear model
FWHM: full width at half maximum
HAMD: Hamilton depression rating scale
HC: Healthy controls
ICD-10: International Statistical Classification of Diseases and Related Health Problems 10th Revision
M1: Primary motor cortex
MADRS: Montgomery Åsberg Depression Rating Scale
MNI: Montreal Neurological Institute
MPRAGE: magnetization prepared rapid gradient echo
PCC: Posterior cingulate cortex
p FWE: family-wise error correction level
p FWEc: cluster-wise correction level
rmANOVA: repeated measures ANOVA
rs-fMRI: Resting state functional MRI
SCL-90-R: Symptom-Checklist-90-R
SMA: Supplementary motor area
SMN: Sensory motor network
UD: Unipolar depressed
UMG: University Medical Center Göttingen
YMRS: Young-Mania-Rating-Scale

## 5. Funding

This work was supported by the University Medical Center Göttingen (UMG) and the German Federal Ministry of Education and Research (Bundesministerium fuer Bildung und Forschung, BMBF: 01 ZX 1507, ‘‘PreNeSt - e:Med’’).

## 6. Competing interests

The authors report no competing interests.

## 7. Acknowledgements

We would like to thank PD Dr. Peter Dechent and Dr. Carsten Schmidt-Samoa for their support and advice.

## Notes

### Competing Interest Statement

The authors have declared no competing interest.

### Funding Statement

This study was funded by Startfoerderung der UMG and by the BMBF.

### Author Declarations

The Ethics committee of the University Medical Center Goettingen (UMG) gave ethical approval for this work.

