## Supplementary material for "Altered functional connectivity relates to motor performance deficits in bipolar but not unipolar depression": Suppl. Material

### Table of contents

1. Materials and methods ... 3

1.1 Medication load ... 3

2. Results ... 7

2.1 Medication ... 7

2.2 Quality check new seed definition technique ... 8

2.2.3 Right hand motor cortex (M1) seeds ... 8

2.2.3 Posterior cingulate cortex (PCC) seeds ... 9

2.3 Frame-by-frame displacements comparison ... 10

2.4 Group and time comparisons SMN-DMN correlation coefficients ... 11

3. References supplementary material ... 13

### 1. Materials and methods

#### 1.1 Medication load

The measurement of medications was performed, whenever possible, in blood and as close as possible to the MRI measurement, but at least 10 hours apart from the last intake to account for first pass metabolization. The reference ranges are given by the central laboratory of the University Medical Center Göttingen (UMG) and complemented by the online version of the Rote Liste (May 2020), if necessary. The medication load was computed as a composite of the sum of ranges,[1–5] varying from below (coded as 1), within (coded as 2) and above (coded as 3) the reference range (Table 1). Exceptions were hormones, excluding Melatonin, and lithium, if blood measurement as part of the study failed, as these are medications individually dosed by the caregivers according to blood levels (coded as 2). Not regularly taking medication or medication that was not considered to be psychoactive (see Table 2), the code was 0.

*Table 1: Medication with reference ranges for medication load*

| **Substance class** | **Substance** | **Metabolite in blood or Rote Liste indication and administration** | **Reference range** | | |
|---|---|---|---|---|---|
| | | | **Min.** | **Max.** | **Unit** |
| Anticholinergic | Biperiden | Pills and retard pills | 3 | 16 | mg |
| Antidepressant | Agomelatine | Pills | 25 | 50 | mg |
| | Amitriptyline | Blood level Amitriptyline + Nortriptyline | 80 | 200 | µg/L |
| | Amitriptyline | Pills and retard pills | 50 | 150 | mg |
| | Bupropion | Pills and retard pills | 150 | 300 | mg |
| | Citalopram | Blood level Citalopram | 50 | 110 | µg/L |
| | Citalopram | Treatment of depression, pills | 20 | 40 | mg |
| | Duloxetine | Blood level Duloxetine | 30 | 120 | µg/L |
| | Duloxetine | Treatment of depression, enteric-coated hard capsule | 60 | 60 | mg |
| | Escitalopram | Blood level Escitalopram | 15 | 80 | µg/L |
| | Escitalopram | Treatment of major depressive disorder, pills | 10 | 20 | mg |
| | Mirtazapine | Blood level Mirtazapine | 30 | 80 | µg/L |
| | Mirtazapine | Pills | 15 | 45 | mg |
| | Sertraline | Blood level Sertraline | 10 | 150 | µg/L |
| | Sertraline | Treatment of depression and obsessive-compulsive disorder, pills | 50 | 250 | mg |
| | Tianeptine | Pills | 37,5 | 37,5 | mg |
| | Trazodone | Outpatient treatment, pills | 200 | 400 | mg |

| Substance class | Substance | Metabolite in blood or Rote Liste indication and administration | Reference range | | |
|---|---|---|---|---|---|
| | | | Min. | Max. | Unit |
| | Venlafaxine | Blood level Venlafaxine + O-Desmethylvenlafaxine | 100 | 400 | µg/L |
| | Venlafaxine | Treatment of major depressive disorder, pills or retard pills | 75 | 375 | mg |
| | Opipramol | Typical dose, pills | 200 | 200 | mg |
| Antihistaminic | Promethazine | Long term treatment, pills | 20 | 100 | mg |
| Anticonvulsive | Gabapentin | Blood level Gabapentin | 2 | 20 | mg/L |
| | Gabapentin | Treatment of epilepsy und peripheral nerve pain, pills or hard capsules | 900 | 3600 | mg |
| | Lamotrigine | Blood level Lamotrigine | 2 | 10 | mg/L |
| | Lamotrigine | Optimal dose bipolar disorder, pills | 200 | 200 | mg |
| | Pregabalin | Blood level Pregabalin | 2 | 5 | mg/L |
| | Pregabalin | Treatment of generalized anxiety disorder, hard capsule | 150 | 600 | mg |
| Benzodiazepine | Alprazolam | Pills | 0,75 | 4 | mg |
| | Diazepam | Outpatient treatment, pills | 5 | 10 | mg |
| | Lorazepam | Sleep disorders due to anxiety and inner tension, pills | 0,5 | 2,5 | mg |
| Dopamin agonists | Rotigotine | Restless-Legs-syndrome transdermal patch, dose per 24h | 1 | 3 | mg |
| Hormones | Dehydro-epiandrosteron | Substitution of hormones | individual | | |
| | Hydrocortisone | Substitution of hormones | individual | | |
| | L-Thyroxin | Substitution of hormones | individual | | |
| | Melatonin | Retard pills | 2 | 2 | mg |
| Mood stabilizer | Lithium | Blood level Lithium | 0,6 | 1,2 | mmol/L |
| | Lithium | Pills | Reference after blood level only | | |
| | Valproate acid | Blood level Valproate acid (amedes MVZ wagnerstibbe für Laboratoriumsmedizin) | 50 | 100 | mg/L |
| | Valproate acid | Blood level Valproate acid (central laboratory UMG) | 346 | 696 | µmol/L |
| | Valproate acid | Bipolar disorder, different galenic | 1000 | 2000 | mg |
| Muscle relaxer | Baclofen | Pills | 30 | 75 | mg |
| Neuroleptic | Aripiprazole | Blood level Aripiprazole | 150 | 500 | µg/L |

| Substance class | Substance | Metabolite in blood or Rote Liste indication and administration | Reference range | | |
|---|---|---|---|---|---|
| | | | Min. | Max. | Unit |
| | Aripiprazole | Manic episodes of Bipolar-I-Disorder, pills | 15 | 30 | mg |
| | Benperidol | Maintenance dose, pills | 1 | 6 | mg |
| | Clozapine | Blood level Clozapine | 350 | 600 | µg/L |
| | Clozapine | Dose range therapy resistant schizophrenia, pills | 200 | 450 | mg |
| | Flupentixol | Maintenance dose, pills | 5 | 20 | mg |
| | Haloperidol | Blood level Haloperidol | 1 | 10 | µg/L |
| | Haloperidol | Treatment of moderate to severe manic episodes in bipolar-I-disorder, drops | 2 | 10 | mg |
| | Olanzapine | Blood level Olanzapine | 20 | 80 | µg/L |
| | Olanzapine | Treatment of bipolar disorder, pills | 5 | 20 | mg |
| | Pipamperone | Minimum for sleep disorder, Maximum general use, pills | 40 | 360 | mg |
| | Quetiapine | Blood level Quetiapine | 100 | 500 | µg/L |
| | Quetiapine | Ad-on-treatment for major depressive disorder, retard pills | 50 | 300 | mg |
| | Quetiapine | Treatment of severe depressive episodes in bipolar disorder, retard pills | 300 | 300 | mg |
| | Quetiapine | Treatment of severe depressive episodes, pills | 300 | 300 | mg |
| | Risperidone | Blood level Risperidone + 9-OH-Risperidone | 20 | 60 | µg/L |
| | Risperidone | Treatment of mania in bipolar disorder, pills | 1 | 6 | mg |
| | Ziprasidone | Acute treatment of schizophrenia and mania in bipolar disorder, hard capsule | 80 | 160 | mg |
| Non-Benzodiazepines | Zolpidem | Pills | 10 | 10 | mg |
| | Zopiclone | Pills | 7,5 | 7,5 | mg |
| Opioid agonists | Oxycodone/ Naloxone | Retard pills | 20/ 10 | 20/ 10 | mg |
| Opioid antagonists | Naltrexone | Typical dose opioid or alcohol addiction, pills | 50 | 50 | mg |

The reference ranges above are provided by the central laboratory of the UMG and complemented by the digital version of Rote Liste (https://www.rote-liste.de/), Liste Arzneimittelinformation für Deutschland).

*Table 2: Medication not considered as psychotropic*

| |
|---|
| Acetylsalicylic |
| Allopurinol |
| Amlodipine |
| Bisoprolol |
| Candesartan |
| Cholecalciferol |
| Diclofenac |
| Dihydralazine |
| Disulfiram |
| Doxazosin |
| Enalapril |
| Etoricoxib |
| Fluvastatin |
| Folic acid |
| Fondaparinux |
| Hydrochlorothiazide |
| Ibuprofen |
| Kalium |
| Lercanidipine |
| Magnesium |
| Metformin |
| Metoprolol |
| Novaminsulfon |
| Omeprazole |
| Pantoprazole |
| Phenprocoumon |
| Ramipril |
| Sitagliptin |
| Spironolactone |
| Torsemide |
| Valsartan |
| Mixture of B-Vitamins |
| Vitamin B1 |

### 2. Results

#### 2.1 Medication

*Table 3: Number of unipolar (UD) and bipolar (BD) depression patients per antidepressant in sessions 1 and 2*

| Substance | UD1 | BD1 | UD2 | BD2 |
|---|---|---|---|---|
| Agomelatine | 2 | 0 | 2 | 0 |
| Amitriptyline | 4 | 2 | 5 | 2 |
| Bupropion | 4 | 0 | 4 | 0 |
| Citalopram | 1 | 2 | 1 | 0 |
| Duloxetine | 6 | 2 | 5 | 2 |
| Escitalopram | 5 | 0 | 3 | 0 |
| Mirtazapine | 9 | 2 | 7 | 1 |
| Sertraline | 12 | 2 | 8 | 2 |
| Tianeptine | 1 | 0 | 1 | 0 |
| Trazodone | 1 | 0 | 1 | 0 |
| Venlafaxine | 6 | 10 | 9 | 9 |
| Opipramol | 1 | 1 | 0 | 0 |

*Table 4: Number of unipolar (UD) and bipolar (BD) depression patients per neuroleptic in sessions 1 and 2*

| Substance | UD1 | BD1 | UD2 | BD2 |
|---|---|---|---|---|
| Aripiprazole | 0 | 3 | 0 | 2 |
| Benperidol | 0 | 2 | 0 | 1 |
| Clozapine | 0 | 2 | 0 | 2 |
| Flupentixol | 0 | 1 | 0 | 1 |
| Haloperidol | 0 | 0 | 0 | 1 |
| Olanzapine | 0 | 4 | 0 | 2 |
| Pipamperone | 0 | 0 | 1 | 0 |
| Quetiapine | 13 | 13 | 11 | 14 |
| Risperidone | 4 | 5 | 5 | 3 |
| Ziprasidone | 0 | 0 | 1 | 0 |

*Table 5: Number of unipolar (UD) and bipolar (BD) depression patients per other psychotropic medication in sessions 1 and 2*

| **Substance class** | **Substance** | **UD1** | **BD1** | **UD2** | **BD2** |
|---|---|---|---|---|---|
| Anticholinergic | Biperiden | 0 | 1 | 0 | 2 |
| Antihistaminic | Promethazine | 1 | 2 | 0 | 2 |
| Anticonvulsive | Gabapentin | 0 | 1 | 0 | 1 |
| | Lamotrigine | 1 | 4 | 2 | 2 |
| | Pregabalin | 2 | 2 | 2 | 1 |
| Benzodiazepine | Alprazolam | 1 | 0 | 0 | 0 |
| | Diazepam | 1 | 3 | 1 | 2 |
| | Lorazepam | 6 | 3 | 4 | 3 |
| Dopamine agonists | Rotigotine | 1 | 0 | 1 | 0 |
| Hormones | Dehydroepi-androsteron | 1 | 0 | 1 | 0 |
| | Hydrocortisone | 1 | 0 | 1 | 0 |
| | L-Thyroxin | 2 | 5 | 2 | 4 |
| | Melatonin | 2 | 1 | 0 | 1 |
| Mood stabilizer | Lithium | 2 | 12 | 3 | 15 |
| | Valproate acid | 0 | 6 | 0 | 7 |
| Muscle relaxer | Baclofen | 0 | 1 | 0 | 0 |
| Non-Benzodiazepines | Zolpidem | 0 | 1 | 0 | 0 |
| | Zopiclone | 1 | 0 | 0 | 0 |
| Opioid/Opioid-antagonists | Oxycodone/ Naloxone | 1 | 0 | 1 | 0 |
| Opioid antagonists | Naltrexone | 0 | 0 | 1 | 0 |

#### 2.2 Quality check new seed definition technique

##### 2.2.3 Right hand motor cortex (M1) seeds

No difference in the volume of individual M1 seeds was seen between groups ($F(2,76) = 0.649$; $p = 0.525$) or across time ($F(1,76) = 0.218$; $p = 0.642$) (Fig. 1, supplementary material). Surprisingly, there was a significant interaction between group and time ($F(2,76) = 3.774$; $p = 0.027$) but since a comparison of the extend of functional changes across the groups was out of the scope of this work this interaction is out of relevance for potential bias. These results remained the same when excluding the lefthanders from analysis.

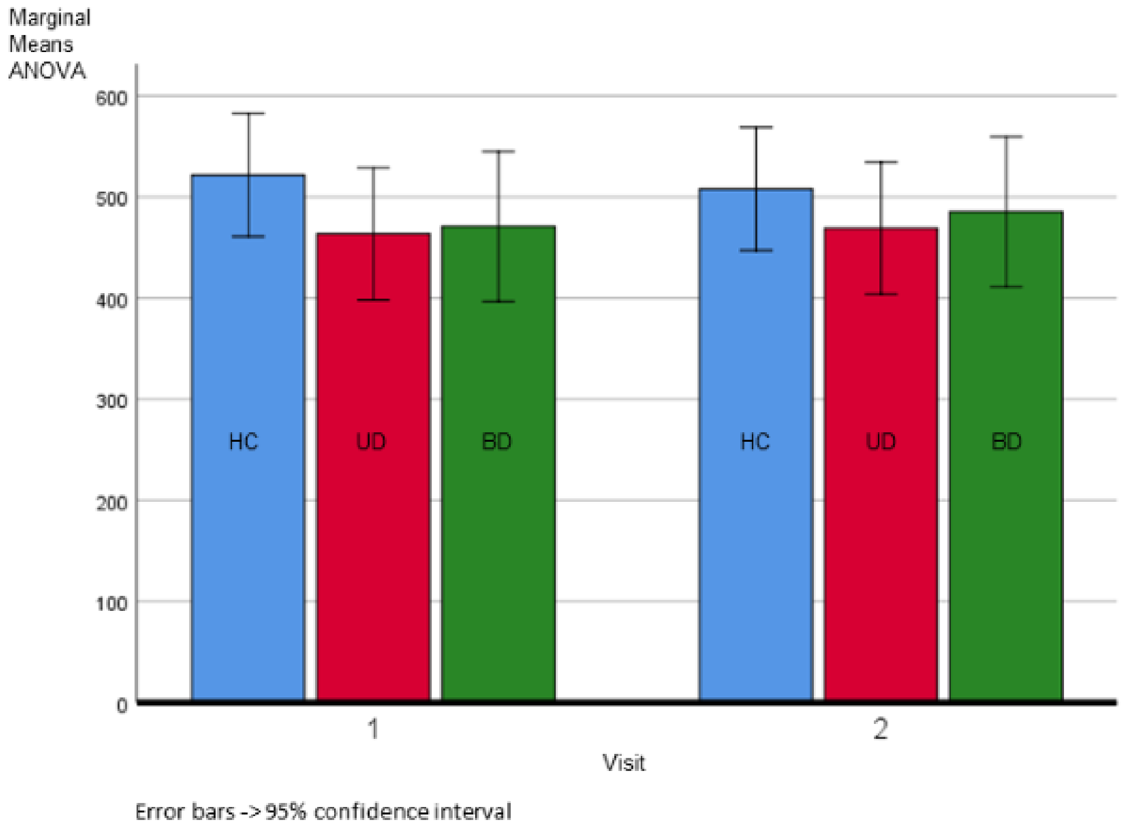


*Figure 1. Marginal means and confidence intervals from repeated-measures ANOVA of right hand motor cortex seed size (in mm³) for healthy controls (HC, blue), unipolar (UD, red) and bipolar (BD, green) patients. HC visit 1 M ± SD = 521.8 ± 139.6 mm³, UD visit 1 M ± SD = 463.4 ± 185.7 mm³, BD visit 1 M ± SD = 470.9 ± 189.6 mm³, HC visit 2 M ± SD = 507.9 ± 128.6 mm³, UD visit 2 M ± SD = 469.0 ± 189.4 mm³, BD visit 2 M ± SD = 485.3 ± 197.5 mm³*

##### 2.2.3 Posterior cingulate cortex (PCC) seeds

No difference in the volume of individual PCC seeds was seen between groups ($F(2,76) = 1.130$; $p = 0.328$) or across time ($F(1,76) = 0.133$; $p = 0.716$) (Fig. 2 supplementary material). Also, there was no interaction seen between group and time ($F(2,76) = 0.886$; $p = 0.417$). These results remained the same when excluding the lefthanders from analysis.

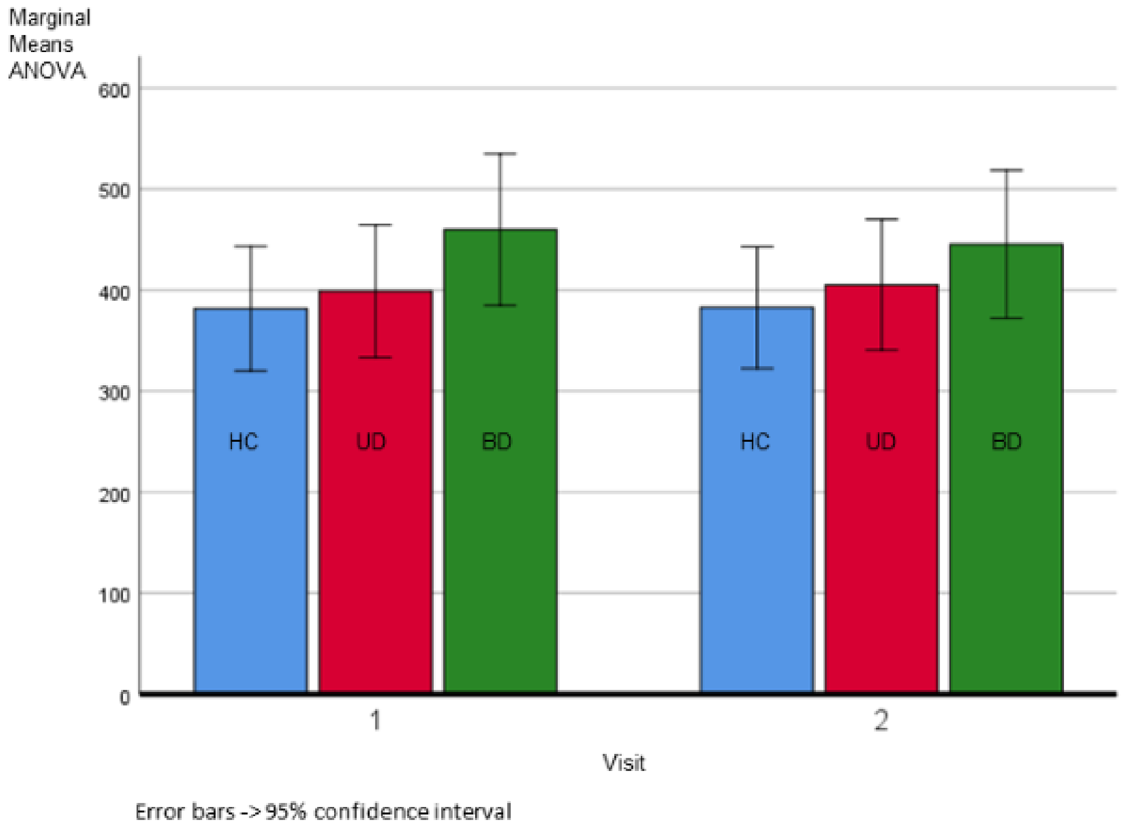


*Figure 2. Marginal means and confidence intervals from repeated-measures ANOVA of posterior cingulate cortex seed size (in $mm^3$) for healthy controls (HC, blue), unipolar (UD, red) and bipolar (BD, green) patients. HC visit 1 M ± SD = 381.4 ± 146.8 $mm^3$, UD visit 1 M ± SD = 398.8 ± 157.7 $mm^3$, BD visit 1 M ± SD = 459.8 ± 219.0 $mm^3$, HC visit 2 M ± SD = 382.5 ± 140.1 $mm^3$, UC visit 2 M ± SD = 405.3 ± 156.0 $mm^3$, BD visit 2 M ± SD = 445.3 ± 215.3 $mm^3$*

#### 2.3 Frame-by-frame displacements comparison

In the rmANOVA of the individual root-mean-squares of the frame-by-frame displacement a difference between groups was seen (F(2,76) = 3.716; p = 0.029) (Fig. 3, supplementary material), although the Games-Howell-post-hoc tests (applied because assumption of homogeneity of variance was not met) did not show a significant difference (HC-UD p = 0.519; HC-BD p = 0.257; UD-BD p = 0.062). There was neither a significant effect of time (F(1,76) = 0.002; p = 0.969) seen nor a group x time interaction (F(2,76) = 0.297; p = 0.744).

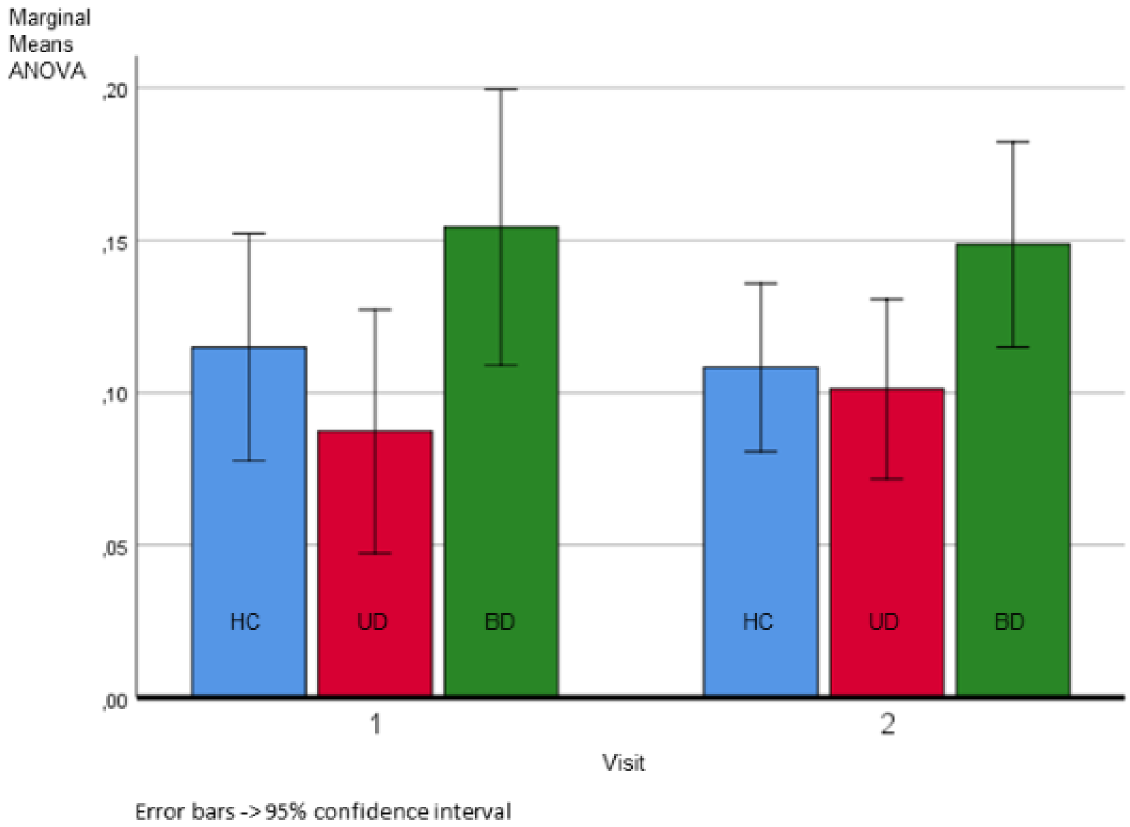


*Figure 3. Marginal means and confidence intervals from repeated-measures ANOVA of root-mean-squares of the frame-by-frame displacement (in mm) for healthy controls (HC, blue), unipolar (UD, red) and bipolar (BD, green) patients. HC visit 1 M ± SD = 0.12 ± 0.08 mm, UD visit 1 M ± SD = 0.09 ± 0.06 mm, BD visit 1 M ± SD = 0.15 ± 0.16 mm, HC visit 2 M ± SD = 0.11 ± 0.07 mm, UC visit 2 M ± SD = 0.10 ± 0.08 mm, BD visit 2 M ± SD = 0.15 ± 0.09 mm*

#### 2.4 Group and time comparisons sensory motor network (SMN) - default mode network (DMN) correlation coefficients

*Table 6: Group and time comparison of SMN-DMN correlation coefficients*

| **HC visit 1 M ± SD** | **UD visit 1 M ± SD** | **BD visit 1 M ± SD** | **HC visit 2 M ± SD** | **UD visit 2 M ± SD** | **BD visit 2 M ± SD** | **rmANOVA factors** |
|---|---|---|---|---|---|---|
| **posterior DMN-SMN (left handers included)** | | | | | | |
| -0.23 ± 0.24 | -0.09 ± 0.25 | -0.10 ± 0.26 | -0.12 ± 0.23 | -0.16 ± 0.19 | 0.01 ± 0.21 | Group: F(2,76) = 3.031, p = 0.054, group effect driven by BD-HC (Post-Hoc p = 0.049) |

| | | | | | | |
|---|---|---|---|---|---|---|
| | | | | | | Time:<br>F(1,76) = 2.328, p = 0.131<br>Group x Time:<br>F(2,76) = 3.232, p = 0.045 |
| **anterior DMN-SMN (left handers included)** | | | | | | |
| -0.01 ± 0.23 | -0.13 ± 0.21 | -0.16 ± 0.21 | -0.08 ± 0.18 | = -0.18 ± 0.18 | 0.01 ± 0.14 | Group:<br>F(2,76) = 3.571, p = 0.033,<br>group effect driven by UD-HC (Post-Hoc p = 0.032)<br>Time:<br>F(1,76) = 0.327, p = 0.569<br>Group x Time:<br>F(2,76) = 6.760, p = 0.002 |
| **DMN-SMN (left handers excluded, gICAs did not show a split of DMN)** | | | | | | |
| -0.16 ± 0.23 | -0.22 ± 0.27 | -0.33 ± 0.16 | -0.10 ± 0.21 | -0.24 ± 0.21 | -0.10 ± 0.20 | Group:<br>F(2,72) = 2.603,<br>p = 0.081<br>Time:<br>F(1,72) = 8.710,<br>p = 0.004<br>Group x Time:<br>F(2,72) = 4.588,<br>p = 0.013 |

*HC = Healthy controls, UD = unipolar group, BD = bipolar group, rmANOVA = repeated measures ANOVA*

### 3. References supplementary material
